# Model-Specific Disruption of Demographic Prediction in Retinal Fundus Images

**DOI:** 10.64898/2026.06.23.26356379

**Authors:** Iyad Majid, Mengyu Wang

## Abstract

**Purpose:** To test whether disease-aware adversarial attacks can disrupt demographic classification in color fundus photographs (CFPs) while retaining glaucoma classification and whether effects transfer across model architectures.

**Methods:** This retrospective study included 13,959 CFPs from 4,271 patients. Vision Transformer (ViT) models classified glaucoma, race, sex, and ethnicity. Standard and disease-aware FGSM, PGD, C&W, and diffusion attacks changed demographic predictions; disease-aware attacks added a glaucoma classification loss term. Fixed ViT-generated perturbations were tested on ResNet50 and EfficientNetB0. Performance was measured using area under the receiver operating characteristic curve (AUC), accuracy, and image-similarity metrics.

**Results:** Baseline glaucoma AUCs were 0.958–0.963 and demographic AUCs were 0.955–0.992. Disease-aware attacks better preserved glaucoma classification while strongly altering demographic predictions. Disease-aware PGD preserved sex-cohort glaucoma AUC at 0.909 (95% CI, 0.894–0.922) while sex AUC fell to 0.000 (95% CI, 0.000–0.000). Disease-aware diffusion preserved ethnicity-cohort glaucoma AUC at 0.946 (95% CI, 0.936–0.956) while ethnicity AUC was 0.001 (95% CI, 0.000–0.002). Demographic changes transferred poorly to ResNet50 and EfficientNetB0.

**Conclusions:** For PGD, C&W, and diffusion, disease-aware attacks retained more glaucoma classification performance than standard attacks while strongly disrupting targeted demographic prediction. Weak cross-architecture transfer indicates model-specific effects rather than demographic information removal.

**Translational Relevance:** Methods intended to reduce sensitive information in retinal images could support multicenter AI development, but failure of one demographic classifier does not establish de-identification. Cross-model evaluation while preserving disease classification may provide a more reliable framework for clinical data sharing.

## Introduction

Glaucoma is a major cause of irreversible blindness, and retinal imaging artificial intelligence (AI) has been studied as a scalable approach to disease detection.^1–18^ Yet model performance can change across institutions, devices, and patient demographic groups.^19–23^ Color fundus photographs (CFPs) also contain demographic and patient-specific information that can be predicted by deep learning models.^24–28^ This creates concerns for sensitive-attribute inference and for disease models that may use demographic or correlated image features during prediction.^29–35^ Methods that can alter demographic prediction while retaining disease-related information may therefore be useful for studying and controlling what retinal AI models learn from images.

Adversarial perturbations offer one image-level approach. Small changes to an image can sharply alter model predictions while leaving much of the visual content unchanged.^36–43^ Unlike methods that modify model training, adversarial perturbations act directly on the image and can target a specific prediction while constraining changes to other outputs. In ophthalmology, prior studies have used adversarial perturbations to test retinal disease-model vulnerability and to explain glaucoma model predictions.^44,45^ These studies focused on altering or explaining disease predictions rather than selectively changing demographic prediction while retaining disease classification.

The idea of selectively attacking images has been explored extensively in face privacy. Chhabra et al. used adversarial perturbations to force incorrect prediction of selected facial attributes while preserving identity and image quality.^46^ FlowSAN similarly attacked face images to confound gender classifiers while retaining face recognition performance and specifically examined whether these effects generalized to unseen classifiers.^47^ These studies show the appeal of adversarial perturbations: unwanted model predictions may be changed without severely degrading the image or modifying its intended use.

However, changing one classifier’s output is not the same as removing the information it predicts. Some face-privacy methods define protection by successful misclassification of the targeted attribute classifier.^46^ For a binary classifier, this can produce an important failure mode: an AUC below 0.5 reflects reversed ranking rather than loss of predictive information, and an AUC of 0 can become 1 when the score direction is reversed.

Generalization is a second concern. FlowSAN was developed in part because protection against one attribute classifier may not extend to unseen classifiers.^47^ Related work outside imaging has reached the same conclusion. Elazar and Goldberg found that demographic information remained recoverable by a new classifier even after the adversarial classifier reached chance performance,^48^ and Kumar et al. showed more broadly that failure of a probing classifier does not establish that the underlying concept was removed.^49^

We tested these issues in retinal imaging. We developed disease-aware attacks that change race, sex, and ethnicity predictions while limiting loss of glaucoma classification performance. We compared FGSM, PGD, C&W, and diffusion attacks in a targeted Vision Transformer (ViT), measured image similarity, and applied the same perturbations to ResNet50 and EfficientNetB0. We asked two questions: whether demographic predictions could be strongly altered while preserving glaucoma classification, and whether those changes persisted across model architectures. This study extends prior ophthalmic adversarial work by targeting demographic prediction while retaining disease classification performance and testing whether these effects persist across architectures.

## Methods

### Dataset and Preprocessing

We retrospectively analyzed 13,959 CFPs from 4,271 patients imaged at Massachusetts Eye and Ear from 2008 through 2020. Available demographic attributes were race, sex, and ethnicity. Glaucoma status was assigned using linked visual field results: images were labeled glaucomatous when mean deviation (MD) was less than −3 dB or the glaucoma hemifield test was abnormal (GHT = 3) with pattern standard deviation (PSD) greater than 1; images were labeled healthy when MD was −3 dB or greater or GHT was normal (GHT = 1) with PSD of 1 or less. This definition identified functional evidence of glaucomatous visual field loss and followed previously used MD, GHT, and PSD criteria.^31,50,51^

### Analysis Cohorts and Experimental Workflow

Because demographic labels were not uniformly available, separate race, sex, and ethnicity cohorts included images with valid glaucoma and corresponding demographic labels. Each cohort was divided at the patient level into stratified training, validation, and test sets (80:10:10; random seed, 42). Images were resized to 224 × 224 pixels and normalized to [0, 1].

For each demographic task, ViT classifiers were trained independently to predict glaucoma and the target demographic attribute from clean CFPs. Baseline performance was measured on held-out test images. A separate perturbation was generated for each test image and demographic task. Standard attacks optimized demographic classification loss alone, whereas disease-aware attacks also penalized loss of glaucoma classification performance. The same targeted ViT classifiers were evaluated on clean and perturbed images. Amplified perturbation maps in **Figure 1** are shown only for visualization and were not used in quantitative analyses.

**Figure 1.**
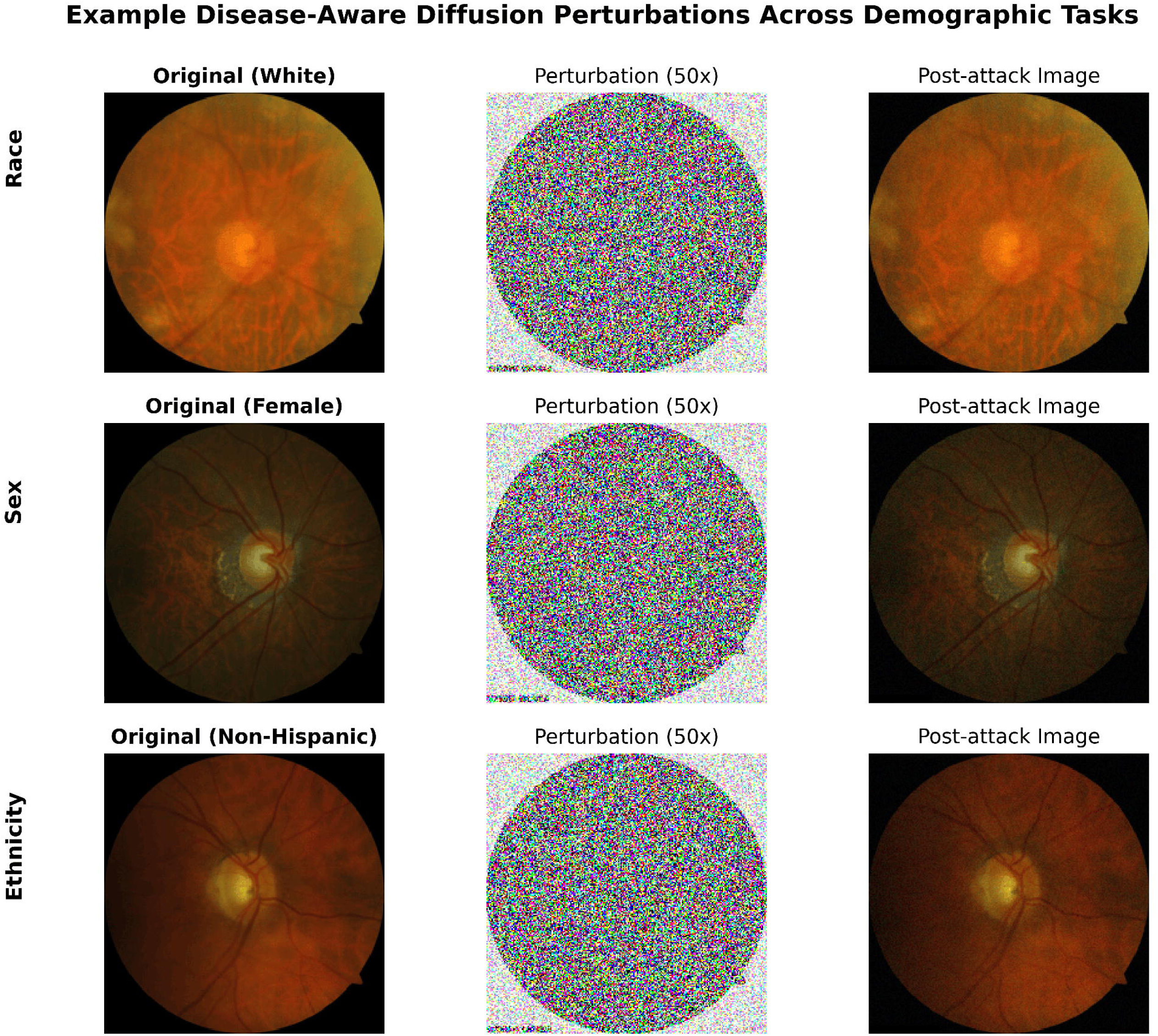
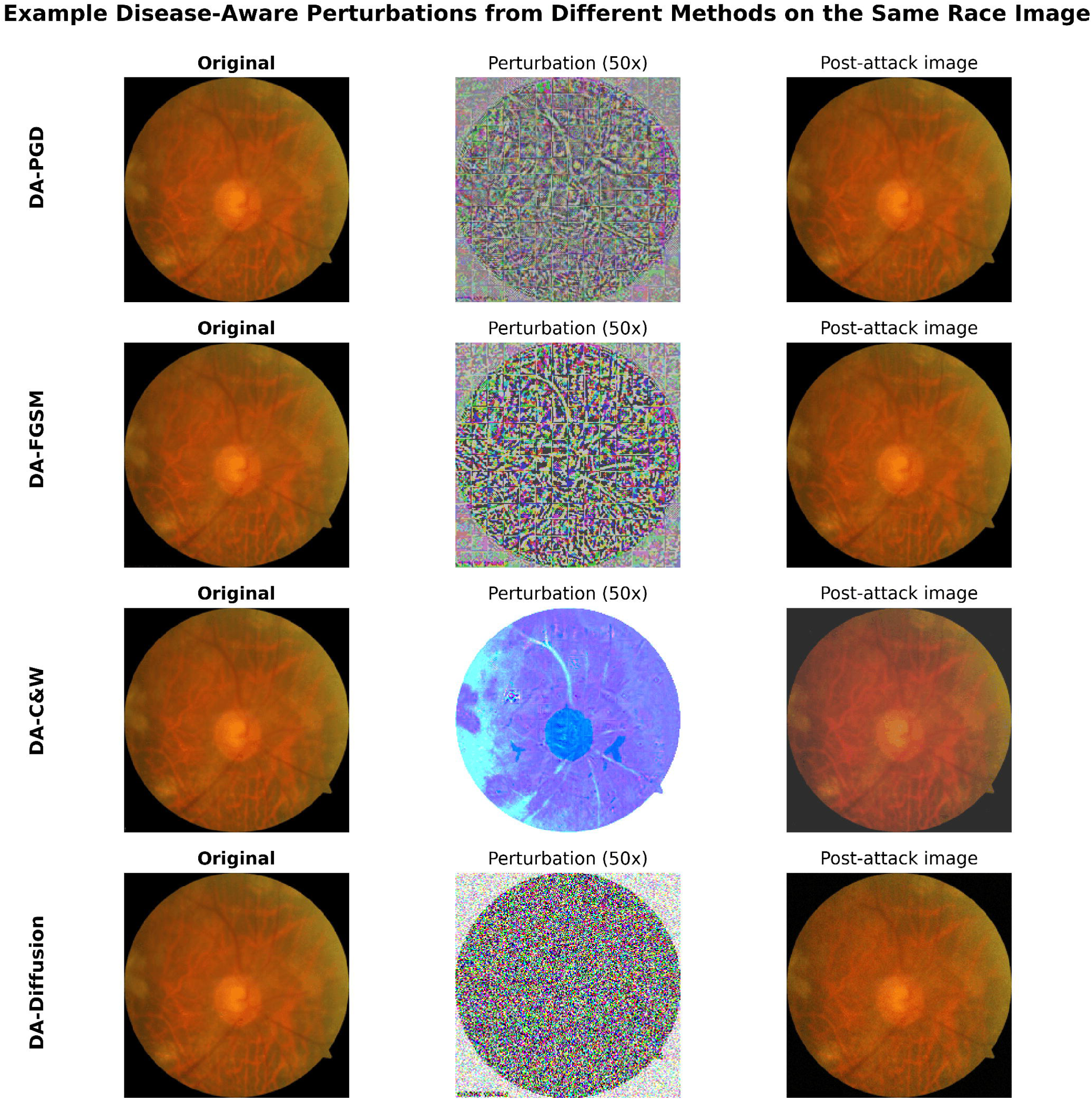
Qualitative examples of disease-aware adversarial perturbations applied to retinal fundus images. (A) Representative disease-aware diffusion perturbations across race, sex, and ethnicity tasks. True demographic labels are shown in parentheses. Perturbation maps are amplified 50× for visualization; post-attack images show the actual perturbed images used for model evaluation. (B) Representative disease-aware perturbations generated by PGD, FGSM, C&W, and diffusion-based attacks on the same race-task fundus image. For each method, the original image, amplified perturbation map, and post-attack image are shown.

For cross-architecture transfer, analogous glaucoma and demographic classifiers were trained using ResNet50 and EfficientNetB0. Fixed perturbations generated with ViT were applied without modification to the same images and evaluated with each unseen architecture.

### Classifier Training

For each architecture, separate classifiers were trained for glaucoma, race, sex, and ethnicity. ViT-B/16, ResNet50, and EfficientNetB0 were initialized with ImageNet-pretrained weights. Models used Adam optimization (learning rate, 1 × 10−4; batch size, 32), a maximum of 50 epochs, and early stopping after 10 epochs without validation improvement. Multiclass race prediction used categorical cross-entropy; binary tasks used binary cross-entropy. Training included horizontal flipping, rotation, zoom, brightness and contrast augmentation, dropout of 0.3, and learning-rate reduction on plateau.

### Disease-Aware Adversarial Attack Formulation

Four attack families were evaluated: single-step FGSM,^39^ iterative PGD,^38^ optimization-based C&W,^37^ and a guided diffusion-based attack.^52,53^ Standard attacks increased demographic classification loss. Disease-aware (DA) attacks added a glaucoma classification term to the same attack-specific objective to decrease demographic classification performance while maintaining glaucoma classification performance.

The disease-aware objective was defined as:

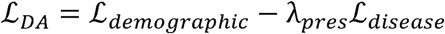

Where *ℒ_demographic_* is demographic classification loss, *ℒ_disease_* is glaucoma classification loss, and *λ_pres_* is a constant that controls the trade-off between demographic prediction disruption and glaucoma classification performance. Perturbations maximized the demographic term while penalizing increases in glaucoma loss. Standard attacks used *λ_pres_* = 0, and DA attacks used *λ_pres_* = 0.9. Final attack parameters were selected on validation data based on demographic prediction change, glaucoma classification performance, and visual plausibility and are provided in **Supplementary Table S1**.

### Image Similarity Analysis

Perceptual similarity between clean and perturbed CFPs was quantified using structural similarity index measure (SSIM), peak signal-to-noise ratio (PSNR), learned perceptual image patch similarity (LPIPS), L∞ perturbation magnitude, and mean absolute perturbation. Metrics were calculated on images scaled to [0, 1], with 95% CIs estimated using 1,000 bootstrap resamples.

### Cross-Architecture Transfer

Cross-architecture transfer was assessed because white-box attacks are optimized using a specific model’s gradients and may not transfer to other models.^54,55^ Glaucoma classification transfer was summarized as the ratio of post-perturbation to baseline glaucoma AUC. Demographic transfer was summarized as the change in demographic AUC.

### Evaluation Metrics and Experimental Setup

The primary metric was area under the receiver operating characteristic curve (AUC), with accuracy as a secondary metric. Glaucoma and demographic performance were compared before and after perturbation. Race performance was summarized using macro-averaged multiclass AUC. Paired P values and 95% CIs were estimated using 1,000 bootstrap resamples. Experiments used TensorFlow 2.14 on NVIDIA RTX A6000 graphics processing units with fixed random seeds.

### Data and Code Availability

The Mass General Brigham Institutional Review Board approved this retrospective study, waived informed consent, and determined that it was conducted in accordance with the Declaration of Helsinki. Code for model training and perturbation generation is available from the corresponding author on request.

## Results

### Cohort Characteristics

The analytic cohort included 13,959 CFPs from 4,271 patients (**Supplementary Table S2**). Mean (SD) age was 63.4 (14.3) years; 53.4% of images were from female patients, and 63.7% had glaucoma. The cohort was 72.1% White, 17.4% Black, and 10.5% Asian; 2.9% of images were from Hispanic patients. The median visual field MD was - 5.0 dB (IQR, −9.8 to −0.4 dB).

### Baseline ViT Performance

Baseline ViT glaucoma AUCs were 0.958 (95% CI, 0.948–0.967), 0.960 (95% CI, 0.951–0.968), and 0.963 (95% CI, 0.955–0.971) in the race, sex, and ethnicity cohorts, respectively (**Figure 2**). Corresponding demographic AUCs were 0.955 (95% CI, 0.945– 0.963), 0.983 (95% CI, 0.977–0.989), and 0.992 (95% CI, 0.987–0.995), showing strong demographic prediction from CFPs.

**Figure 2.**
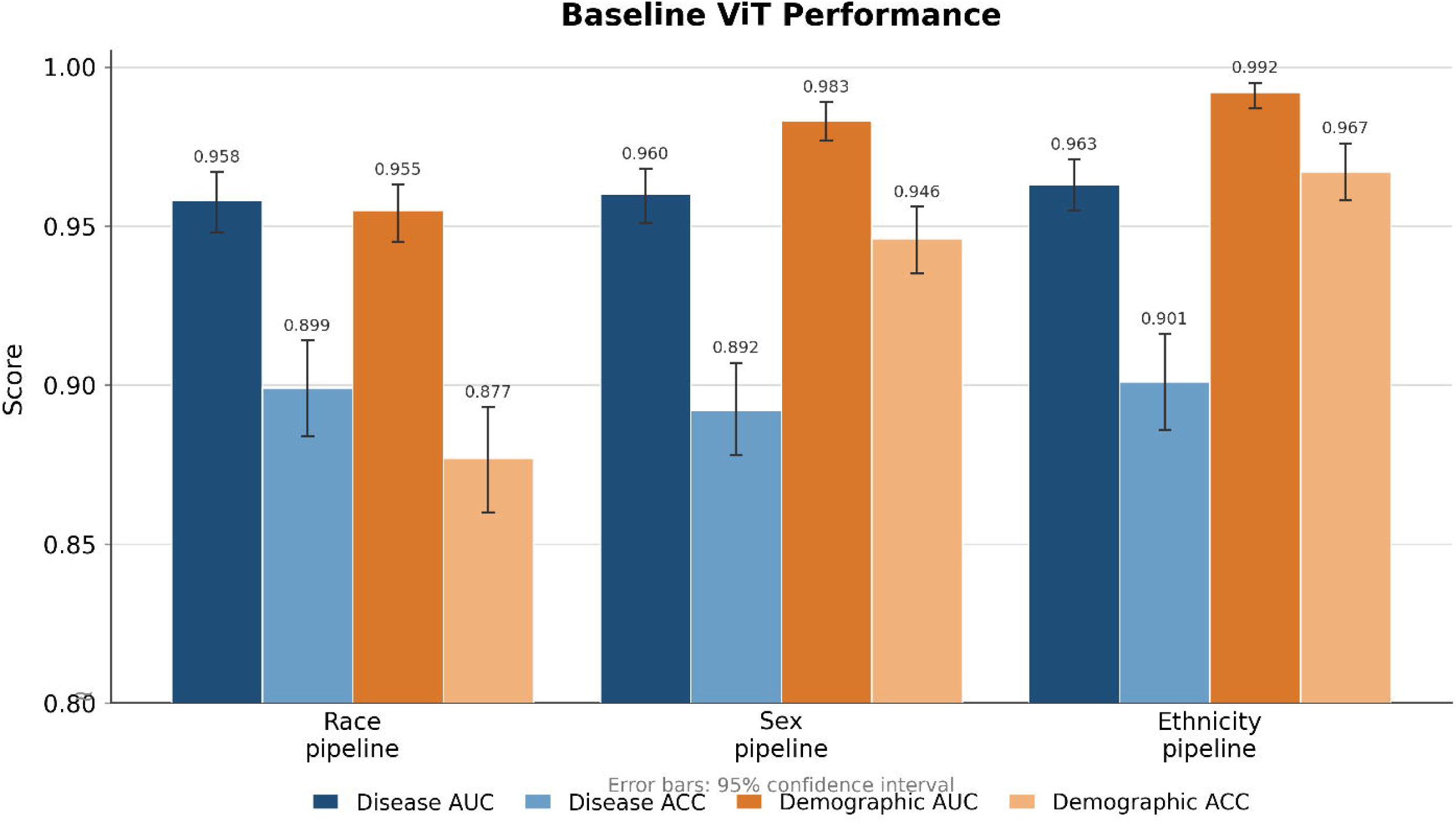
Baseline ViT Performance. Baseline Vision Transformer performance for glaucoma and demographic prediction across race, sex, and ethnicity cohorts. Bars show AUC and accuracy; error bars indicate 95% bootstrap confidence intervals. AUC = area under the receiver operating characteristic curve; ViT = Vision Transformer.

### PGD and FGSM Attacks

All reported baseline-to-attack comparisons were statistically significant (P < .01). Standard attacks strongly changed demographic predictions but generally caused larger losses in glaucoma classification performance. Disease-aware attacks retained more glaucoma classification performance for PGD, C&W, and diffusion (**Figure 3**).

**Figure 3.**
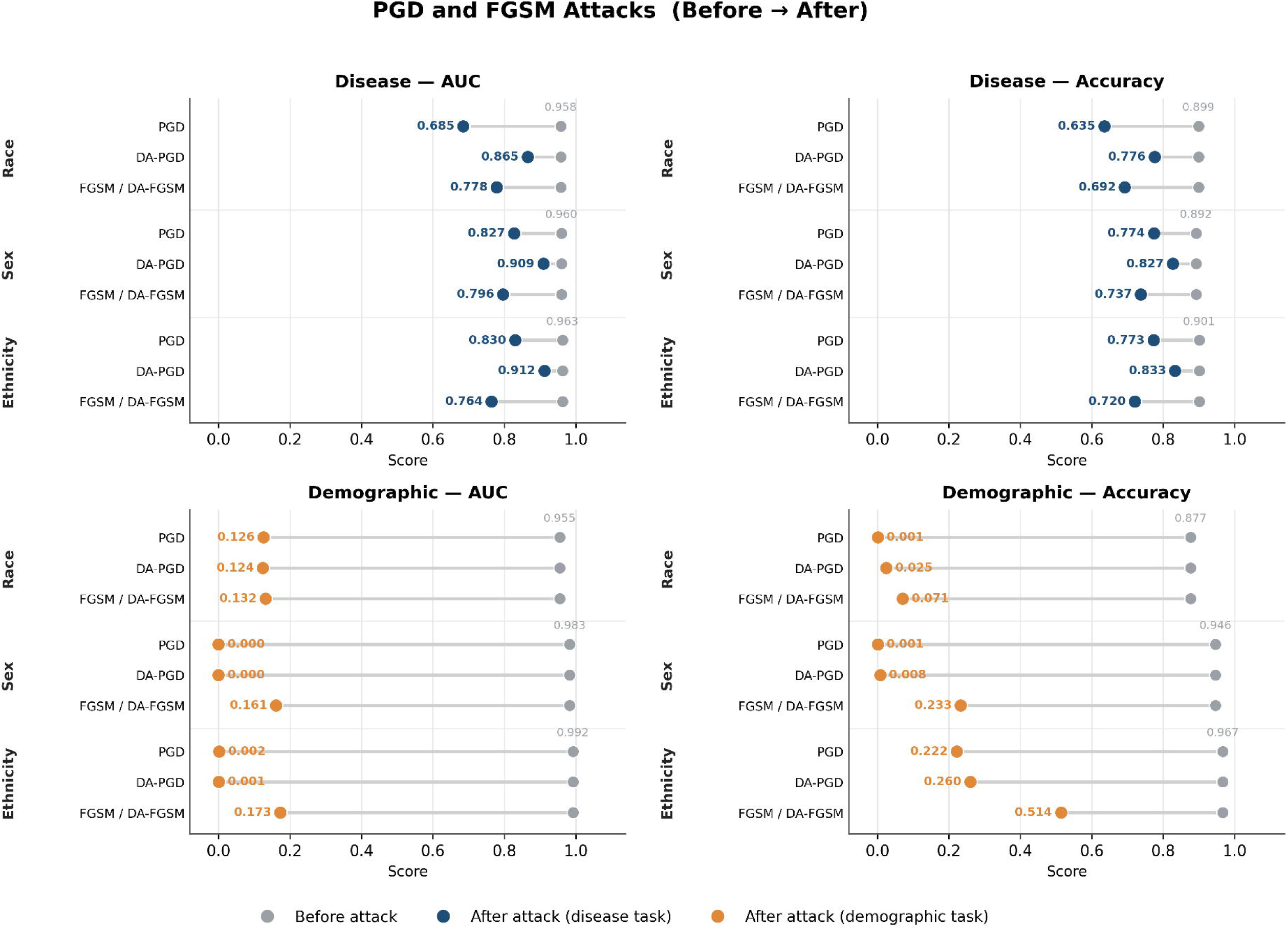
PGD and FGSM Attacks. Effect of standard and disease-aware PGD and FGSM attacks on glaucoma and demographic classification performance. Points show performance before and after perturbation across race, sex, and ethnicity tasks for glaucoma AUC, glaucoma accuracy, demographic AUC, and demographic accuracy. AUC = area under the receiver operating characteristic curve; DA = disease-aware; FGSM = Fast Gradient Sign Method; PGD = Projected Gradient Descent.

With standard PGD, demographic AUCs fell to 0.126 for race, 0.000 for sex, and 0.002 for ethnicity, while glaucoma AUCs decreased to 0.685, 0.827, and 0.830, respectively. DA-PGD retained glaucoma AUCs of 0.865 (95% CI, 0.845–0.883), 0.909 (95% CI, 0.894–0.922), and 0.912 (95% CI, 0.897–0.925), while demographic AUCs were 0.124, 0.000, and 0.001. FGSM produced demographic AUCs of 0.132 to 0.173 and glaucoma AUCs of 0.764 to 0.796; its disease-aware variant produced nearly identical results.

### C&W and Diffusion Attacks

Across PGD, C&W, and diffusion attacks, sex and ethnicity AUCs were often near 0 (**Figure 4**). Standard C&W produced demographic AUCs of 0.303 for race, 0.001 for sex, and 0.006 for ethnicity but reduced glaucoma AUCs to 0.655, 0.744, and 0.686. DA-C&W increased glaucoma AUCs to 0.931 (95% CI, 0.919–0.943), 0.940 (95% CI, 0.929–0.951), and 0.935 (95% CI, 0.923–0.945), with demographic AUCs of 0.322, 0.000, and 0.020. Standard diffusion attacks produced glaucoma AUCs of 0.756 to 0.862 and demographic AUCs of 0.000 to 0.213. DA-Diffusion retained glaucoma AUCs of 0.926 (95% CI, 0.910–0.941), 0.927 (95% CI, 0.915–0.939), and 0.946 (95% CI, 0.936–0.956), with demographic AUCs of 0.230, 0.006, and 0.001 for race, sex, and ethnicity.

**Figure 4.**
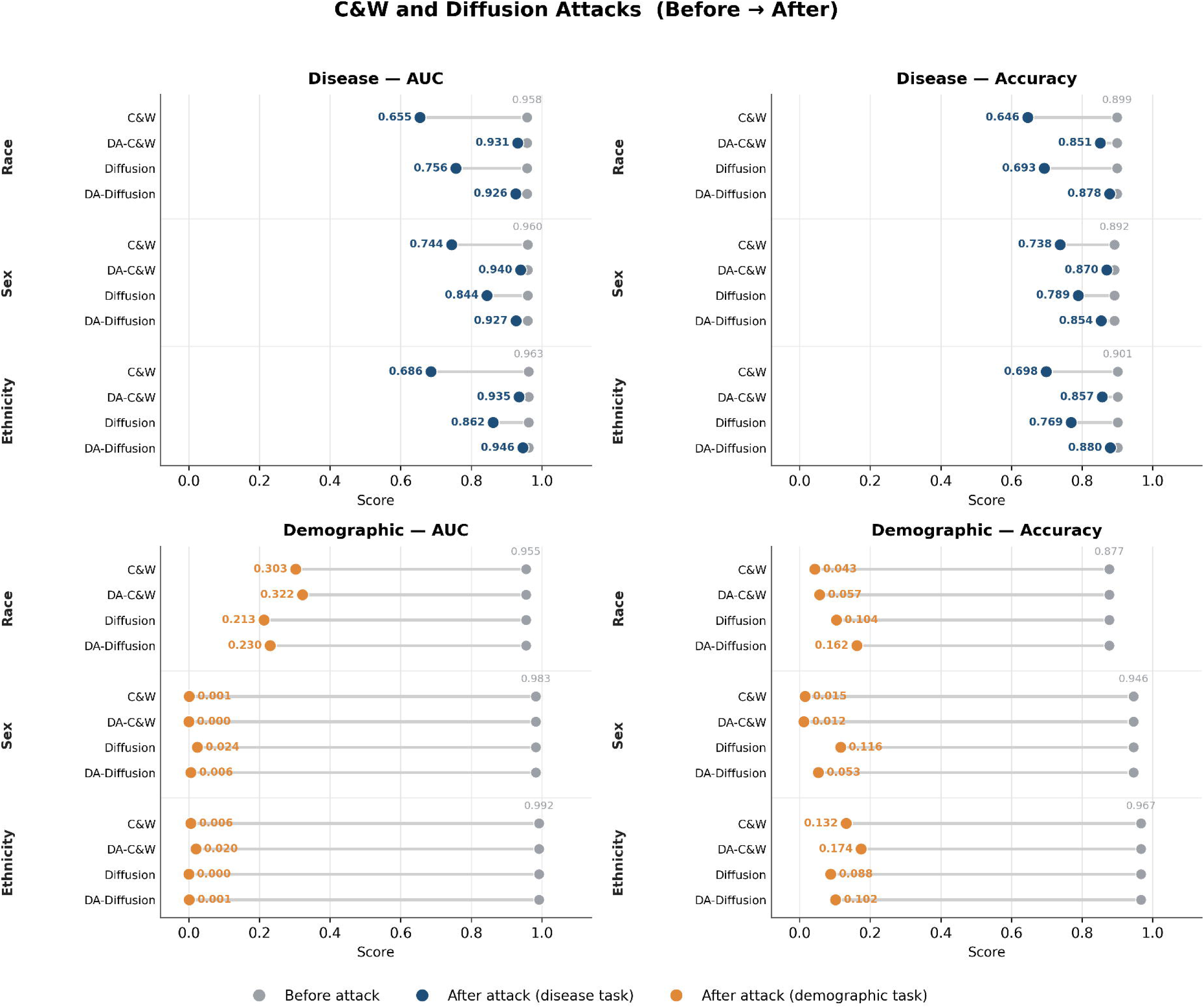
C&W and Diffusion Attacks. Effect of standard and disease-aware C&W and diffusion attacks on glaucoma and demographic classification performance. Points show performance before and after perturbation across race, sex, and ethnicity tasks for glaucoma AUC, glaucoma accuracy, demographic AUC, and demographic accuracy. AUC = area under the receiver operating characteristic curve; C&W = Carlini & Wagner; DA = disease-aware.

### Image Similarity

Image similarity differed across attack families (**Supplementary Table S3**). PGD provided the highest image similarity, while diffusion retained more glaucoma classification performance with larger image changes.

DA-PGD had the highest similarity to the original images, with mean SSIM of 0.991 to 0.993, PSNR of 51.2 to 52.8 dB, LPIPS of 0.003 to 0.005, and L∞ magnitude of 0.005.

DA-FGSM also produced small changes (SSIM, approximately 0.974; PSNR, approximately 46.5 dB; LPIPS, approximately 0.019; L∞ magnitude, 0.005).

DA-Diffusion produced larger changes (SSIM, 0.693–0.694; PSNR, approximately 34.1 dB; LPIPS, 0.177–0.180; L∞ magnitude, 0.030), and DA-C&W produced the largest changes (SSIM, 0.672–0.674; PSNR, approximately 19.3 dB; LPIPS, 0.233–0.240; L∞ magnitude, 0.217–0.243).

### Cross-Architecture Transfer

ViT-generated perturbations showed weak cross-architecture transfer for demographic prediction on ResNet50 and EfficientNetB0 (**Table 1**). Glaucoma classification performance transferred more consistently than the demographic prediction changes.

**Table 1.** Best Cross-Architecture Transfer Results. Best transferred demographic suppression for each demographic attribute and target architecture. Values show the attack with the greatest demographic AUC reduction for each setting. Disease AUC ratio represents post-perturbation disease AUC divided by baseline disease AUC.

| Demographic | Target model | Best transferred attack | Disease AUC preservation ratio (95% CI) | Demographic AUC reduction (95% CI) |
| --- | --- | --- | --- | --- |
| Race | ResNet50 | DA-Diffusion | 0.978 (0.971–0.985) | 0.021 (0.016–0.028) |
| Race | EfficientNetB0 | C&W | 0.863 (0.845–0.881) | 0.090 (0.078–0.104) |
| Sex | ResNet50 | DA-C&W | 0.915 (0.903–0.928) | 0.047 (0.038–0.056) |
| Sex | EfficientNetB0 | DA-Diffusion | 0.842 (0.822–0.861) | 0.294 (0.270–0.318) |
| Ethnicity | ResNet50 | DA-C&W | 0.904 (0.888–0.919) | 0.038 (0.025–0.053) |
| Ethnicity | EfficientNetB0 | Diffusion | 0.848 (0.828–0.867) | 0.293 (0.247–0.336) |

For race, the largest AUC decrease on ResNet50 was 0.021 with DA-Diffusion (glaucoma AUC ratio, 0.978), whereas C&W decreased race AUC by 0.090 on EfficientNetB0 but lowered the glaucoma AUC ratio to 0.863. For sex, DA-C&W decreased AUC by 0.047 on ResNet50 (glaucoma AUC ratio, 0.915), and DA-Diffusion decreased AUC by 0.294 on EfficientNetB0 (glaucoma AUC ratio, 0.842). For ethnicity, DA-C&W decreased AUC by 0.038 on ResNet50 (glaucoma AUC ratio, 0.904), and diffusion decreased AUC by 0.293 on EfficientNetB0 (glaucoma AUC ratio, 0.848). PGD and FGSM caused little demographic prediction change on the unseen architectures; C&W and diffusion caused larger changes but more often reduced glaucoma classification performance.

## Discussion

Three findings define this study. First, for PGD, C&W, and diffusion, disease-aware attacks retained more glaucoma classification performance than standard attacks while strongly changing demographic predictions in the targeted ViT. Second, sex and ethnicity AUCs often fell near 0, showing classifier inversion rather than loss of demographic information. Third, demographic prediction changes transferred poorly to ResNet50 and EfficientNetB0. Together, these results show that failure of a targeted demographic classifier does not mean the demographic information has been removed from a CFP.

Classifier inversion matters because it can look like strong protection if lower AUC is always treated as better. Prior face-privacy studies defined protection by flipping gender predictions or forcing attribute classifiers to misclassify.^46,56^ Chhabra et al. explicitly reported flipped score distributions while describing the selected attributes as anonymized.^46^ Later work showed that targeted perturbations could fail on unseen classifiers,^47^ and studies of adversarial representation removal found that demographic or concept information could remain after the training adversary or probe was defeated.^48,49^ Our retinal results show the same measurement problem: below-chance AUC reflects a changed decision rule, not proof that the information is absent.

The cross-architecture results make this distinction clear. ViT-generated perturbations caused large demographic prediction changes in ViT but much smaller changes in ResNet50 and EfficientNetB0. This is consistent with the model-specific nature of adversarial perturbations and prior work showing that attribute protection can fail on unseen classifiers.^47,54,55^ Cross-architecture transfer is therefore an important test of whether an effect extends beyond the targeted model, although transfer alone does not establish that the underlying sensitive information is irrecoverable.

Recent work has similarly applied adversarial perturbations to retinal image privacy. Ghaffary and Ebrahimi Moghaddam developed a knowledge-distillation framework that perturbed fundus images to disrupt patient identity recognition while preserving retinopathy classification, reporting high identity fooling rates across multiple unseen models.^57^ Their findings support the feasibility of selectively altering privacy-sensitive retinal features while retaining disease-model utility. However, identity misclassification and de-identification are not equivalent: a high fooling rate demonstrates that recognition models assign incorrect identities but does not establish that patient-specific information is no longer recoverable by a newly trained classifier or other inference strategy. Their evaluation also differs from ours in targeting biometric identity rather than demographic attributes and does not address the below-chance AUC inversion observed in binary demographic classification. Our results therefore extend this work by showing that apparent suppression itself requires careful interpretation: even profound deterioration of a targeted classifier may reflect model-specific disruption or reversed prediction rather than removal of the underlying sensitive information.

Disease-aware optimization still provided a useful tradeoff. Compared with standard PGD, C&W, and diffusion attacks, the disease-aware variants generally retained more glaucoma classification performance while producing large changes in targeted demographic prediction. PGD also produced the highest image similarity. These results show that the disease-aware objective can preserve model utility while changing targeted outputs, even when it does not remove the underlying demographic information.

This study used one retinal dataset and fixed demographic classifiers; new classifiers were not trained on the perturbed images. Glaucoma classification performance and image-similarity metrics also do not establish unchanged human interpretation. These limits do not change the main finding that targeted demographic prediction disruption was strongly model-dependent.

The practical implication is simple: demographic AUC from the attacked model should not be used alone as evidence of demographic information removal. Evaluation should distinguish chance-level prediction from classifier inversion and test whether the effect holds in other models.

In conclusion, for PGD, C&W, and diffusion, disease-aware attacks retained more glaucoma classification performance than standard attacks while strongly disrupting targeted demographic prediction. Near-zero binary AUCs reflected classifier inversion, and weak cross-architecture transfer showed that the effect was model-specific. Fooling one demographic classifier is not the same as removing demographic information from the image.

## Data Availability

The dataset used in this study is institutionally restricted due to patient privacy considerations. Code for all experiments, including model training and adversarial attack generation, is available from the corresponding author upon request.

## Meeting Presentation

Presented in part as a poster at the Association for Research in Vision and Ophthalmology Annual Meeting, Denver, Colorado, May 2026.

## Financial Support

This work was supported by the National Institutes of Health, Bethesda, Maryland, United States, grants R01 EY036222, R21 EY035298, and P30 EY003790. The sponsor or funding organization had no role in the design or conduct of this research.

## Financial Relationship(s) Disclosure

Iyad Majid: no commercial relationship. Mengyu Wang: no commercial relationship.

## Conflict of Interest

No conflicting relationship exists for any author.

## Notes

### Competing Interest Statement

The authors have declared no competing interest.

### Author Declarations

No human subjects were included in this study. This retrospective study was approved by the Mass General Brigham Institutional Review Board (IRB) and conducted in accordance with the principles outlined in the Declaration of Helsinki. The IRB also deemed informed consent to be waived for this study.

### Summary of Updates

This version has been revised to clarify the interpretation and generalizability of adversarial demographic prediction disruption in retinal fundus images. We added cross architecture evaluation using ResNet50 and EfficientNetB0 to determine whether perturbations generated for a Vision Transformer generalized to unseen model architectures. We also revised the interpretation of very low binary demographic AUC values, emphasizing that near zero AUC can reflect classifier inversion rather than removal of demographic information. The Introduction and Discussion were updated to distinguish targeted classifier failure from true de identification and to place the findings in the context of prior adversarial privacy and concept removal literature. We added discussion of recent retinal privacy work using adversarial perturbations for patient identity protection and highlighted differences in privacy endpoints and evaluation. The manuscript was also revised for clearer terminology, more concise framing, updated references, and improved consistency throughout.

