## Supplementary Table S1 for "Model-Specific Disruption of Demographic Prediction in Retinal Fundus Images"

**Supplementary Table S1. Attack hyperparameters used for standard and disease-aware adversarial perturbation experiments.** DA attacks incorporated a disease-preservation term with $\lambda = 0.9$, whereas standard attacks used only the demographic attack objective. FGSM, PGD, and diffusion-based attacks were constrained with $\varepsilon = 0.005$. C&W was optimized with an L2 penalty and box constraints but without an explicit $L_\infty$ bound. DA = disease-aware; FGSM = Fast Gradient Sign Method; PGD = Projected Gradient Descent; C&W = Carlini & Wagner.

| **Attack** | **Variants evaluated** | **Perturbation bound** | **Steps / iterations** | **Step size / learning rate** | **Disease-preservation weight** | **Additional parameters** | **Pixel clipping** |
|---|---|---|---|---|---|---|---|
| FGSM | Standard, DA-FGSM | $\varepsilon = 0.005$ | 1 gradient step | Step size = $\varepsilon$ | $\lambda = 0.9$ for DA; $\lambda = 0$ for standard | Entropy weight = 0.0 | Clipped to [0, 1] |
| PGD | Standard, DA-PGD | $\varepsilon = 0.005$ | 5 PGD steps | $\varepsilon / 5 = 0.001$ | $\lambda = 0.9$ for DA; $\lambda = 0$ for standard | Entropy weight = 0.0 | Clipped to [0, 1] |
| C&W | Standard, DA-C&W | No explicit $L_\infty$ bound; box-constrained to valid image range | 100 optimization iterations | Adam learning rate = 0.01 | $\lambda = 0.9$ for DA; $\lambda = 0$ for standard | $c = 1.0$; $\kappa = 0.0$; L2 penalty | Clipped to [0, 1] |
| Diffusion | Standard, DA-Diffusion | $\varepsilon = 0.005$ | 50 diffusion-guidance steps | Noise scale = 0.01; guided step size = 0.01 / (t + 1) | $\lambda = 0.9$ for DA; $\lambda = 0$ for standard | Gaussian noise added at each step | Clipped to [0, 1] |
| General settings | All attacks | Images normalized to [0, 1] | — | — | — | Batch size = 16; image size = 224 × 224 | Clipped to [0, 1] |
