## Supplementary Table S2 for "Model-Specific Disruption of Demographic Prediction in Retinal Fundus Images"

**Supplementary Table S2. Cohort characteristics.** Values are reported at the image level unless otherwise specified. MD = mean deviation; PSD = pattern standard deviation; GHT = glaucoma hemifield test; IQR = interquartile range; SD = standard deviation.

| Characteristic | N = Images |
|---|---|
| Images, n | 13,959 |
| Unique patients, n | 4,271 |
| Laterality code 0, n (%) | 6,797 (48.7%) |
| Laterality code 1, n (%) | 7,162 (51.3%) |
| Healthy, n (%) | 5,074 (36.3%) |
| Glaucoma, n (%) | 8,885 (63.7%) |
| Race: Asian, n (%) | 1,469 (10.5%) |
| Race: Black, n (%) | 2,424 (17.4%) |
| Race: White, n (%) | 10,066 (72.1%) |
| Sex: Female, n (%) | 7,449 (53.4%) |
| Sex: Male, n (%) | 6,510 (46.6%) |
| Ethnicity: Non-Hispanic, n (%) | 13,551 (97.1%) |
| Ethnicity: Hispanic, n (%) | 408 (2.9%) |
| Age, mean ± SD, years | 63.4 ± 14.3 |
| Age, median [IQR], years | 65.0 [55.2, 73.4] |
| Visual field MD, mean ± SD, dB | −6.3 ± 6.7 |
| Visual field MD, median [IQR], dB | −5.0 [−9.8, −0.4] |
| Visual field PSD, mean ± SD | 2.9 ± 2.3 |
| Visual field PSD, median [IQR] | 4.0 [0.0, 5.0] |
| GHT code 1, n (%) | 5,074 (36.3%) |
| GHT code 3, n (%) | 8,885 (63.7%) |
