## Supplementary Table S3 for "Model-Specific Disruption of Demographic Prediction in Retinal Fundus Images"

**Supplementary Table S3. Quantitative Imperceptibility of Disease-Aware Adversarial Perturbations.** Mean SSIM, PSNR, LPIPS, L∞ magnitude, and mean absolute perturbation for disease-aware attacks across demographic tasks. Higher SSIM/PSNR and lower LPIPS/perturbation values indicate greater visual similarity.

| DA attack | SSIM, mean range | PSNR, mean range | LPIPS, mean range | L∞ mean | Mean abs perturbation |
|---|---|---|---|---|---|
| DA-PGD | 0.991–0.993 | 51.2–52.8 | 0.003–0.005 | 0.005 | 0.0019–0.0023 |
| DA-FGSM | 0.974–0.975 | 46.5 | 0.019 | 0.005 | 0.0045 |
| DA-Diffusion | 0.693–0.694 | 34.1 | 0.177–0.180 | 0.030 | 0.0170 |
| DA-C&W | 0.672–0.674 | 19.3 | 0.233–0.240 | 0.217–0.243 | 0.0855–0.0862 |
